# Vegetable intake and cardiovascular risk: genetic evidence from Mendelian randomization

**DOI:** 10.1101/2022.03.21.22272719

**Authors:** Qi Feng, Andrew J. Grant, Qian Yang, Stephen Burgess, Jelena Bešević, Megan Conroy, Wemimo Omiyale, Yangbo Sun, Naomi Allen, Ben Lacey

## Abstract

**Background:** Observational studies have demonstrated inverse associations between vegetable intake and cardiovascular diseases. However, the results are prone to residual confounding. The separate effects of cooked and raw vegetable intake remain unclear. This study aims to investigate the association between cooked and raw vegetable intake with cardiovascular outcomes using Mendelian randomization (MR).

**Methods:** We identified 15 and 28 genetic variants associated statistically and biologically with cooked and raw vegetable intake, which were used as instrumental variables to estimate the associations with coronary heart disease (CHD), stroke, heart failure (HF) and atrial fibrillation (AF). In one-sample analysis using individual participant data from UK Biobank, we adopted two stage least square approach. In two-sample analysis, we used summary level statistics from genome-wide association analyses. The independent effects of cooked and raw vegetable intake were examined with multivariable MR analysis. The one-sample and two-sample estimates were combined via meta-analysis. Bonferroni correction was applied for multiple comparison.

**Results:** In the meta-analysis of 1.2 million participants on average, we found null evidence for associations between cooked and raw vegetable intake with CHD, HF, or AF. Raw vegetable intake was nominally associated with stroke (odds ratio [95% confidence interval] 0.82 [0.69 – 0.98] per 1 serving increase daily, p = 0.03), but this association did not pass corrected significance level.

**Conclusions:** Cooked and raw vegetable intake was not associated with CHD, AF or HF. Raw vegetable intake is likely to reduce risk of stroke, but warrants more research. Solely increasing vegetable intake may have limited protection, if any, on cardiovascular health. This calls for more rigorous assessment on health burden associated with low vegetable consumption.

## 1. Introduction

Cardiovascular diseases (CVDs) are the leading cause of global burden of disease [1] and are caused by a complex interplay of genetic and environmental factors [2,3]. It is estimated that 8 million CVD deaths and 188 million CVD disability-adjusted life years are attributable to unhealthy diet annually [3], of which up to 20% may be due to insufficient vegetable intake [4].

There exists a substantial body of observational evidence to support an inverse association between vegetable intake and CVDs [5,6], which has led to international dietary guidelines that recommend higher intake of vegetable for primary prevention [7–9]. Indeed, a meta-analysis of 45 cohort studies found that higher vegetable intake level was associated with 13% lower risk of CVD [10], with other meta-analyses reporting similar risk reductions for coronary heart disease (CHD) and stroke [11]. In spite of large sample size, long follow-up time and adjustment for multiple confounders in some of the studies [12–14], the observed associations may be biased by residual confounding, as vegetable intake tends to be correlated with socioeconomic status and other lifestyle factors [15]. A previous study estimated that a large proportion of the observed association was actually accounted for by residual confounding [16]. In addition, most studies have examined vegetable intake as a whole and the separate effects of raw and cooked vegetable intakes remain inconsistent [12,14,17].

Mendelian randomization (MR) is an epidemiological design using genetic variants, usually single nucleotide polymorphisms (SNPs), as instrumental variables for causal inference. Conceptually similar to a randomized controlled trial in the way that genetic variants are randomly assorted at meiosis and fixed after fertilization, MR is able to limit the potential bias of residual confounding and reverse causality, thus making causal inference possible [18]. Therefore, the objective of this study was to investigate the effects of cooked and raw vegetable intakes on cardiovascular risk in a multivariable MR design.

## 2. Methods

In this study, we performed both two-sample MR and one-sample MR to quantify the associations of cooked and raw vegetable intakes with cardiovascular outcomes. Two-sample MR was performed with summary level statistics of genome-wide association studies (GWAS), and one-sample MR with individual-level data of UK Biobank participants. Two-sample and one-sample MR estimates were meta-analysed to obtain overall effect estimates. In both two-sample and one-sample MR, we performed multivariable analysis as the primary analysis, in which cooked and raw vegetable intakes were adjusted for each other, aiming to examine their separate effects. Univariable analysis, in which the effects of cooked and raw vegetable intake were fitted separately, was performed as secondary analysis. An overview of the methods is shown in Figure 1.

### 2.1 Genetic instrument selection

Genetic instruments associated with cooked and raw vegetable intake were identified from three GWAS of UK Biobank [19–21] (Supplementary table 1). Individual intake of cooked and raw vegetables (in number of heaped tablespoons; one heaped tablespoon is roughly equivalent to one serving) were measured with a touchscreen questionnaire on diet frequency at recruitment in UK Biobank. The repeatability and validity of the diet questionnaire has been evaluated and confirmed in a previous analysis: the repeatability was 82% for cooked vegetable and 72% for raw vegetable when compared to a repeat assessment after four years, and high agreement was observed when compared to 24-hour diet recall assessment[22].

We combined all SNPs that were significant at genome-wide significance level (p < 5 * 10^−8^) in the three GWAS [19–21], and removed those that were duplicated, or rare variants (minor allele frequency < 1%) or in linkage disequilibrium (r^2^ 0.001 and distance 10 000 kb). To further reduce horizontal pleiotropy, we searched the associated phenotypes for each SNP in PhenoScanner v2 database (http://www.phenoscanner.medschl.cam.ac.uk/), and further removed SNPs that were associated with potential confounders, such as smoking, alcohol drinking, blood pressure and adiposity.

In total, we identified 15 and 28 eligible SNPs for cooked and raw vegetable intakes, which explained 0.8% and 2.4% of phenotypic variance, respectively. The SNPs are located in different gene loci. The majority of the loci have expressions in tissues of gastrointestinal tract and/or other organs of the digestive system (Supplementary table 2). The biological mechanisms behind the selected SNPs and vegetable consumption were suggested to be mediated by individual’s taste and smell preference, as some hit SNPs (for example, *rs9323534*) were associated with olfactory receptors [20]. The mechanisms are possibly additionally mediated by their expression and/or regulation on lipid metabolism (*rs17714824, rs33947258, rs12190945, rs78940216, rs17075255, rs11608727*), protein and/or glucose metabolism (*rs6975898, rs11209780, rs57221424, rs6079589*). SNPs for raw vegetable intake were specifically associated with gastrointestinal diseases (*rs11125813* for Crohn’s disease, and *rs4281874* for gastrointestinal dismay); while one SNP for cooked vegetable intake was associated with teeth development (*rs10161952*). All these may contribute to individual’s dietary preference and eventually influence food choices. (Supplementary table 2)

The magnitude of the association between the SNPs and vegetable intakes were extracted from the GWAS by Canela-Xandri et al [21], as this GWAS has a large sample size and was adjusted for more covariates. It was performed in 452264 unrelated European-ancestry individuals, and adjusted for sex, age, age square, array batch, assessment centre and the leading 20 genetic principle components. The strength of the genetic instruments was evaluated with F statistics, with F statistics > 10 suggesting good instrument strength [23]. The process of SNP selection and the characteristics of these SNPs are shown in Supplementary figure 1 and Table 1.

### 2.2 One-sample MR

#### 2.2.1 Data source

We used individual-level data of UK Biobank participants for one sample MR. UK Biobank is a population-based prospective cohort, which recruited half million participants aged 40-69 years between 2006 and 2010 across England, Wales and Scotland [24]. At baseline, participants completed a touchscreen questionnaire that collected information on socioeconomic status, health status, medication use, family disease history, lifestyle, and environmental exposures. Anthropometric and physical measures were taken using standard procedures; blood, urine and saliva samples were collected [24]. Genotypes in UK Biobank were assayed using Affymetrix UKBiLEVE Axiom array® for about 50,000 participants and the UK Biobank Axiom array® for about 440,000 participants. Genetic pre-imputation quality control (QC), phasing, and imputation of genetic data in UK Biobank have been described elsewhere [25]. UK Biobank was approved by the North West Multicentre Research Ethics Committee, the National Information Governance Board for Health and Social Care in England and Wales, and the Community Health Index Advisory Group in Scotland. All participants provided informed consent.

In this analysis, we excluded participants if they (1) did not have individual genotype array data, (2) withdrew from the cohort, (3) did not pass genetic QC, or (4) did not have vegetable intake data. In genetic QC, we excluded participants if (1) the self-reported sex was different from the genetic sex, (2) the sex chromosome karyotypes are putatively different from XX or XY, (3) outliers in heterozygosity and missing rates, indicating the sample genotypes are of poor quality, (4) non-European genetic ethnicity, and (5) genetic relatedness with other participants in UK Biobank. (Supplementary figure 2)

The health status of participants was followed-up via linkage to national death registries (NHS Digital for participants in England and Wales; and NHS Central Registry for participants in Scotland) and hospitalization databases (the National Health Service [NHS] Hospital Episode Statistics for participants in England; the Scottish Morbidity Record for participants in Scotland; and the Patient Episode Database for participants in Wales). At the time of this study, the death registries capture records through 28^th^ February 2021, and the hospitalization databases through 31^st^ March 2021 for participants from England and Scotland, and 28^th^ February 2018 for participants from Wales. Diagnosis of cardiovascular outcomes were ascertained by mapping relevant International Classification of Disease (ICD) version 9 and version 10 codes in death registry (including both primary and secondary causes of death) and hospitalization records (including both primary and secondary diagnosis). We used the following ICD 10 codes: *I21-I25* for CHD; *I60-I61, I63-I64* for stroke; *I63-I64* for ischaemic stroke; *I50, I11*.*0, I13*.*0, I13*.*2* for heart failure (HF); and I48 for atrial fibrillation (AF). The equivalent ICD-9 codes used are shown in Supplementary table 3.

#### 2.2.2 Statistical analysis

Unweighted genetic risk scores (GRS) for cooked and raw vegetable intakes were calculated by summing the number of vegetable intake-increasing alleles a participant carried and dividing it by the total number of SNPs. We estimated the association between GRS and baseline characteristics of the population, by fitting linear regressions of the baseline characteristics (age, sex, body mass index, physical activity, alcohol drinking, smoking, systolic blood pressure, diastolic blood pressure, red meat intake, processed meat intake and oily fish intake) on GRS, and using the p value for the overall model fitting as the p value for potential associations.

MR estimates were obtained with two-stage least square method, in which two regressions were fitted. In the first stage, we fitted a multivariate linear regression model of the two GRS on cooked and raw vegetable intake, adjusted for sex, age, age square, assessment centre, genotype batch and the first 20 genetic principal components, in participants without cardiovascular disease (non-cases). From this first stage regression, we obtained the genetically predicted cooked and raw vegetable intakes. In the second stage, we fitted a logistic regression model of the genetically predicted cooked and raw vegetable intakes on the outcome, adjusted for the same covariates as the first stage regression. The covariate adjustment strategy was recommended to reduce potential collider bias [26] and has been used previously [27].

For sensitivity analysis, we fitted a Cox model in the second stage after excluding participants with CVD at recruitment. We also performed sensitivity analysis by additionally adjusting for potential confounders that were found to be associated with the GRS in our analysis (oily fish intake). More details are shown in Supplementary methods.

As secondary analysis, we performed univariable one-sample MR analysis for cooked and raw vegetable intake separately, using a similar two stage least square method. In the first stage, we fitted a linear model for vegetable intake, and obtained the genetically predicted values; in the second stage, a logistic regression of the predicted vegetable intake was fitted on the outcomes. The same covariates were adjusted as in multivariable analysis.

### 2.3 Two-sample MR

#### 2.3.1 Data source

For two-sample MR, we used summary level GWAS statistics of *CARDIoGRAMplusC4D* consortium for CHD (with 60801 cases) [28], *MEGASTROKE* consortium for stroke and ischaemic stroke (with 40585 and 34217 cases, respectively) [29], *HERMES* for heart failure (with 47309 cases) [30], and *Nielson 2018* for atrial fibrillation (with 60662 cases) [31]. For replication, we used the summary level GWAS data of the five cardiovascular outcomes from the *FinnGen* consortium (release 5) [32], using the following FinnGen endpoint codes: “*I9_CHD*” for CHD, “*I9_STR_SAH*” for stroke, “*I9_STR_EXH*” for ischemic stroke, “*I9_HEARTFAIL_NS*” for HF, and “*I9_AF*” for AF, respectively. All these GWAS were performed in unrelated predominantly European-ancestry individuals. *CARDIoGRAMplusC4D* consortium, *MEGASTROKE* and *FinnGen* had no sample overlap with UK Biobank, while *HERMES* and *Nielson 2018* had 40% and 38% sample overlap with UK Biobank, respectively. Basic characteristics of these GWAS are shown in Supplementary table 1.

#### 2.3.2 Statistical analysis

In multivariable two-sample MR, we included the 43 (= 15 + 28) SNPs but further removed three duplicate SNPs. The remaining 40 SNPs are not in linkage disequilibrium. The associations of each SNP with cooked vegetable intake and raw vegetable intake were extracted from the GWAS by Canela-Xandri et al [21] (Supplementary table 4), while the association with the outcomes were extracted from the relevant outcome GWAS data. For the SNPs that could not be matched in outcome GWAS, we first tried to identify proper proxy SNPs in linkage disequilibrium (r^2^ > 0.80, distance < 500 kb); if no proper proxy was identified, the unmatched SNPs were removed from further analysis. Finally, 39 SNPs were used in analysis with FinnGen derived GWAS data (*rs11608727* was not matched); otherwise, all 40 SNPs were used.

Summary-level association statistics for each SNP were orientated across different GWAS so that their effect estimates were aligned on the same allele [33]. Inverse variance weighted method was performed to estimate the association between vegetable intake and the outcomes [34].

As secondary analysis, we performed univariable two-sample MR, in which 15 and 28 SNPs were used for cooked and raw vegetable intake, respectively. We used inverse-variance weighted method, while sensitivity analyses were performed using alternative approaches, including weighted median method and MR-Egger method. The weighted median method can generate reliable effect estimates when at least 50% of SNPs are valid instrument [35]. MR-Egger method can detect and correct for possible directional pleiotropy [35]. Pleiotropy was examined with MR-Egger intercept test, with a p value < 0.05 suggesting presence of directional pleiotropy, in which case MR-PRESSO method [36] was used to examine the effect of pleiotropy. MR-PRESSO method can detect outlier SNPs and provide effect estimates after removing the outliers.

### 2.4 Meta-analysis

We performed meta-analysis by combining the two-sample and one-sample MR estimates, for univariable MR and multivariable MR separately. Random-effects model was used as the primary analysis, while fixed-effect model was used as sensitivity analysis. I^2^ statistics were calculated to quantify heterogeneity, with I^2^ > 50% indicating presence of high heterogeneity. Since HERMES and Nielson 2018 had sample overlap with UK Biobank, and one-sample MR estimate tends to overestimate the association [37], we performed a sensitivity meta-analysis by excluding the one-sample MR estimates.

The effects were quantified with odds ratio (OR) and its 95% confidence interval (CI), reflecting risk change of the outcome for lifelong increase of one serving of daily vegetable intake. Bonferroni correction was applied to control multiple comparison, α = 0.05/5 = 0.01. The statistical tests were two-sided, with p value < 0.01 as conservative level of statistical significance, and p value between 0.01 and 0.05 as suggestive evidence. All analyses were performed in R environment (version 4.1.1), with “*MendelianRandomization*” package (version 0.5.1), “*MR-PRESSO*” package (version 1.0) and “*meta*” package (version 5.0-1).

## 3. Results

The average F statistics were 29 (range 18 to 48) for the SNPs associated with cooked vegetable intake, and 30 (range 18 and 46) for the SNPs associated with raw vegetable intake, respectively, suggesting good instrument strength. (Table 1)

### 3.1 One-sample MR

In one-sample MR, 361 797 UK Biobank participants were included, with 37 014 cases of CHD, 9 298 stroke, 7264 ischaemic stroke, 11 773 HF, and 25915 AF during 12.1 years of follow-up. The mean age was 56.9 (standard deviation (SD) 7.9) years and 55.0% were women. The mean values of cooked and raw vegetable intake were 2.74 (1.77) and 2.19 (1.98) heaped tablespoons per day, respectively (Supplementary table 5). The correlation between cooked and raw vegetable intake was low (Pearson correlation coefficient = 0.30). The mean GRS for cooked and raw vegetable intakes was 1.10 (0.16) and 1.06 (0.12), respectively (Supplementary figure 3). GRS were strongly associated with actual observed vegetable intake (p < 2*10^−16^), and not associated with age, sex, body mass index, physical activity, smoking, drinking, blood pressure, red meat intake or processed meat intake (Supplementary table 6). However, GRS were positively associated with oily fish intake. The mean F statistics for the cooked vegetable intake GRS and the raw vegetable intake GRS was 67 (range 62 to 70) and 314 (range 289 to 322), respectively (Supplementary table 7).

In multivariable one-sample analysis we did not find significant evidence for associations between vegetable intake and cardiovascular outcomes (Figure 2). Univariable analyses and subsequent sensitivity analyses, including the ones that additionally adjusted for oily fish intake, also generated non-significant associations (supplementary table 7 and 8).

### 3.2 Two sample MR

In multivariable two-sample MR analysis mutually adjusting for the two kinds of vegetable intake, we found null evidence for associations between raw vegetable intake and CHD, stroke, ischemic stroke and HF, consistent across different data sources (Figure 2). The univariable analysis showed similarly null evidence for most of the associations (Supplementary table 9).

Potential presence of directional pleiotropy was found in cooked vegetable intake and ischaemic stroke in *FinnGen* (p for MR-Egger intercept = 0.05) and the association between raw vegetable intake and AF in *FinnGen* (p for MR-Egger intercept = 0.01). However, MR-PRESSO analysis detected zero and one outlier SNP (*rs62380935*), respectively, and removing the outlier yielded very similar results. Weighted median method generated consistent results to inverse-variance weighted estimates. (Supplementary table 9).

### 3.3 Meta-analysis

Combining two-sample and one-sample multivariable MR estimates in meta-analysis, we found suggestive evidence for an inverse association between raw vegetable intake and stroke (0.82 (0.69 – 0.98), p = 0.03), although it failed to pass the Bonforreni corrected significance level (0.01). The associations of raw vegetable intake with other CVD outcomes were directionally inverse (except atrial fibrillation), but none of them were statistically significant: CHD (0.86 (0.72 – 1.02), p = 0.08), ischaemic stroke (0.85 (0.71 – 1.03), p = 0.10) and heart failure (0.88 (0.73 – 1.05), p = 0.15) (Figure 2). Sensitivity analysis by excluding the one-sample estimate in UK Biobank consistently showed suggestive evidence for raw vegetable intake and stroke risk (0.79 (0.66 – 0.95), p = 0.01) (Supplementary table 10). Meta-analysis of univariable estimates showed insignificant, trend for raw vegetable intake with the outcomes (Supplementary table 11, supplementary figure 4). We did not find evidence for associations between cooked vegetable intake and cardiovascular outcomes. There was no evidence of heterogeneity in meta-analysis; fixed-effect model and random-effects model produced similar results.

## 4. Discussion

In this MR meta-analysis of about 1.2 million participants on average, we examined the MR association between cooked and raw vegetable intake with multiple cardiovascular outcomes. The results demonstrated generally null evidence for the effects of vegetable intake on CHD, HF or AF, although we observed suggestive evidence for an inverse association between raw vegetable intake and stroke. Previous meta-analyses of cohort studies have found that higher vegetable intake were associated with reduced CVD risks [10,11]. However, causal inference has been difficult, because the bias of residual confounding is ubiquitous in observational research, while randomized controlled trials with large sample size and long follow-up time for capturing clinical outcomes have been lacking [38,39].

Our findings of null associations, although in contrast to the observational evidence, are supported by recent MR studies [40–42]. Dietary-derived antioxidants, especially vitamin C, vitamin E, retinol, carotene and lycopene, are valid biomarkers reflecting vegetable consumption level [43], and have been proposed as the major mechanisms for the observational inverse association with cardiovascular diseases [44]. Kobylecki et al. used one SNP *rs33972313* in *SLC23A1* gene region, which codes sodium-dependent vitamin C transporter 1, as the genetic instrument for serum vitamin C, and reported that vitamin C was not associated with incident CHD or all-cause mortality in a cohort of 100 000 Danish participants [42]. Zhu et al., using 9 SNPs for serum circulating vitamin C, further found null genetic association, with a range of cardiovascular risk factors and diseases, including CHD, stroke, heart failure, atrial fibrillation, blood pressure, obesity and serum lipids [40]. Luo et al. investigated five antioxidants, i.e., vitamin C, vitamin E, retinol, carotene and lycopene, in both absolute circulating levels and relative metabolite levels, and found null evidence for any of their associations with incident CHD [41]. Similarly, Martens et al. found the five antioxidants were not associated with stroke [45].

Residual confounding bias is likely to be one of the major reasons for the discrepancies between observational and genetic evidence [16,46]. A previous analysis of 400 000 UK Biobank participants [16] estimated that residual confounding accounted for about 80-90% of the observed associations between vegetable intake and CVD outcomes, and this percentage is likely to be higher providing adjustment for unobserved confounders and/or more accurate measurement of the confounders. This is consistent with the findings of this study using MR design. MR is considered to be able to methodologically further limit residual confounding, compared with observational designs [18], which is one of the strengths of our study.

The validity of MR estimates is dependent on three assumptions. First, the instrumental variables are associated with at least one of the exposure variables. Second, the instrumental variables are not associated with confounders of all exposure-outcome associations. Third, there are no independent pathways between the genetic instruments and outcome other than through one or more of the exposures [47]. For the first assumption, we selected the SNPs that are associated with cooked or raw vegetable intake at genome-wide association level from three GWAS. The GRS in one-sample MR are also highly associated with the observed phenotypes (p < 2*10^−16^). Although the explained phenotypic variance was small (0.8% for cooked vegetable intake and 2.4% for raw vegetable intake), which is common for behaviour phenotypes, the high F statistics indicated them as strong instruments. Additionally, the selected SNPs are biologically associated with vegetable intake via their regulations on olfactory receptor, gastrointestinal health, teeth health and metabolism of lipids/protein/glucose. For the second assumption, we searched the Phenoscanner v2 database for any phenotypes associated with the SNPs, and excluded the SNPs that are associated with confounding factors in the association between vegetable intake and CVD, including alcohol drinking, smoking, blood pressure and adiposity. In one-sample MR, the GRS were not associated with common cardiovascular risk factors, such as alcohol drinking, smoking, physical activity, blood pressure, obesity, red meat intake and processed meat intake. Although the GRS were associated with oily fish intake, adjustment for it in the two stage least square analysis did not change the results. For the third assumption, the MR-Egger intercept test did not show strong evidence of directional pleiotropy for most of the analyses; MR-PRESSO analysis generated similar results to the primary inverse-variance weighted estimates. Sensitivity analysis of median method and MR-Egger method also generated consistent results. Therefore, the three assumptions are plausibly satisfied in our study.

Extra caution should be taken when interpreting the findings. MR estimates reflect the risk change in the outcomes for solely increasing one serving of vegetable intake lifelong, while all other risk factors remain unchanged. First, this is different from the interpretation of estimates from randomized controlled trials, which manifest the risk change for an increase of exposure for a temporary period of time. The null associations in our MR analysis suggest that a lifelong exposure of one-serving higher vegetable intake may not decrease CVD risks. Second, it is assumed that all other risk factors for CVD are fixed, including socioeconomic, lifestyle and dietary factors among others. However, diet is always complex, characterised with intake of many different kinds of food and the substitutions between them, where, given the relative stability of an individual’s calorie intake, high consumption of vegetable is associated with lower intake of other food [48,49]. General population should be cautious what kinds of food be replaced, but this is beyond the scope of our study and warrants future research on dietary patterns, which describes the overall diet. In this sense, our findings are consistent with the recommendation of a balanced diet. Our study also calls for the attention to reassess the disease burdens associated insufficient vegetable intake.

We observed a potential inverse association between raw vegetable intake and incident stroke, which passed the conventional significance level (0.05) but failed in Bonferroni corrected significance level (0.01). This is in line with a previous observational study of 20 000 individuals [50]. If this association was a true effect, this may indicate the different health effects of cooked and raw vegetable on stroke risk, which has been suggested by previous observational studies [12,14]. Nevertheless, this remains unclear due to residual confounding and requires future research.

The strengths of this study included MR design to limit residual confounding, large sample size for well powered analysis, carefully selected SNPs that are statistically and biologically associated with vegetable intake, and coherent results from various sensitivity analysis. Nevertheless, this study has some limitations. First, the biological mechanism behind the SNPs and vegetable-eating behaviour is not completely understood. A previous GWAS study of dietary intake showed that vegetable intake was associated with olfactory receptor loci [20]. We additionally found that some of the selected SNPs are associated with metabolism of lipids, glucose and protein, teeth health and gastrointestinal diseases, which are also likely to influence individuals’ dietary preference or choice of vegetables over meat, and cooked vegetable over raw, or vice versa.

Second, although we differentiated cooked and raw vegetable intake in this study, these phenotypes are still a mix of different vegetable kinds and cooking methods (for cooked vegetable). It may be valuable that future studies can further differentiate vegetable kinds and cooking methods. Third, the dietary intake was measured in UK biobank, a cohort based on England, Wales and Scotland, so the findings may be more generalizable to populations that consume similar types of vegetables (e.g., carrots, broccoli, spinach, peppers) and use similar cooking methods. Fourth, vegetable intake was measured with self-reporting questionnaire in UK Biobank. It was not directly validated against valid biomarkers, although comparison to 24-hour recall assessment showed good agreement; nevertheless, the findings are consistent with recent MR studies using antioxidants as biomarkers for vegetable intake.

Fifth, our analysis was confined to a population of European ancestry, which reduced population stratification bias, but may limit its generalizability to populations of other ethnicities. Sixth, the effect estimate in this study may not directly comparable to other studies, as vegetable intake was quantified with the number of tablespoons per day in UK Biobank, instead of grams per day as usually used in other studies. Seventh, there are sample overlaps between UK Biobank with HERMES consortium (40%) and Nielson 2018 (38%), which may overestimate the associations with heart failure and atrial fibrillation; however, we still observed null associations on them, which indicates that the true effect, if any, should be minor.

## Conclusion

We performed this MR analyses and their meta-analysis of large sample size, and found null evidence for association between cooked and raw vegetable intake with CHD, AF and HF. This suggested that solely increasing vegetable intake while keeping intakes of other food unchanged is very unlikely to decrease cardiovascular risk, and calls for reassessment of disease burden of low intake of vegetable. The dietary pattern was measured in British population, which may limit the generalizability of the findings to other populations with different dietary patterns.

## Data Availability

All data produced in the present study are available upon reasonable request to the authors

## Acknowledgement

The funders had no role in the design and conduct of the study; collection, management, analysis, and interpretation of the data; preparation, review, or approval of the manuscript; or the decision to submit the manuscript for publication.

We thank the investigators and participants in CARDIoGRAMplusC4D consortium, MEGASTROKE consortium, HERMES consortium and Nielson 2018 study. We thank the investigators and participants in UK Biobank and FinnGen. Use of individual level data of UK Biobank participants is approved under the application (No. 41115).

## Data availability

The summary level GWAS data can be downloaded at http://www.cardiogramplusc4d.org/steering-committee/ for CARDIoGRAMplusC4D consortium, https://www.megastroke.org/ for MEGASTROKE consortium, https://cvd.hugeamp.org/downloads.html#summary for HERMES consortium. FinnGen summary level GWAS data can be available at https://www.finngen.fi/fi upon application. UK Biobank individual level data is available at https://www.ukbiobank.ac.uk/ upon application. Analytic R codes are available upon request.

